# A Prospective Clinical Evaluation of a Patient Isolation Hood During the COVID-19 Pandemic

**DOI:** 10.1101/2021.02.19.21251739

**Authors:** Forbes McGain, Samantha Bates, Jung Hoon Lee, Patrick Timms, Marion A Kainer, Craig French, Jason Monty

**Author notes:** Corresponding Author: Forbes McGain, Depts. of Anaesthesia and Intensive Care, Footscray Hospital, Gordon St., Footscray, Vic., 3011. **Ethical and TGA approval.** Ethical approval was obtained from Melbourne Health Human Research Ethics Committee (MH HREC 2020.129). The isolation hood was registered with the Therapeutic Goods Administration (TGA) (CT-2020-CTN-01390-1). **COI:** A patent has been filed for the personal ventilation hood by the University of Melbourne/Western Health. The lead authors (Forbes McGain, Jason Monty) were the leads in this patent application. All other authors have no conflicts of interest. **CRediT Statement:** FM conceived the project, wrote the background, methods and ethics submission, sought funding, assisted in data capture, and wrote and revised the manuscript. SB assisted in methods and ethics preparation, ran the data capture and management, and assisted in manuscript preparation. JHL assisted in data capture and trial management, and manuscript preparation. PT assisted in data capture and management, and manuscript preparation. MK assisted in the methods and manuscript preparation. CF assisted in manuscript preparation. JM co-conceived the project, sought funding, and assisted in manuscript preparation.

## Abstract

**Background:** Healthcare workers have frequently become infected with SARS-CoV-2 whilst treating patients with COVID-19. A variety of novel devices have been proposed to reduce COVID-19 cross contamination.

**Objective:** To test whether a novel patient isolation hood was safe and comfortable, and could potentially reduce HCW COVID-19 infections.

**Methods:** Prospective cohort study of 20 patients, entailing staff/patient questionnaires, and safety aspects of prototype isolation hoods.Prospective collection of HCW COVID-19 data.Assessment of the hood’s safety and practicality, and adverse event reporting.

**Outcome Measures:** Questionnaires’ responses, adverse events reporting, rates of HCW infections during study period (20/6/2020 −21/7/2020).HCW COVID-19 infections reported until last recorded HCW COVID-19 diagnosis (20/6/2020 −27/9/2020).

**Results:** Of the 60 (of 64) eligible individual staff surveys, 60 favoured isolation hood use.Staff were unanimous in: perceiving the hood as safe (60/60), preferring its use (56/56), and understanding its potential COVID-19 cross-contamination minimisation (60/60). All eight patients who completed the questionnaire thought the isolation hood helped prevent COVID-19 cross-infection, was safe, and comfortable. There were no reported patient safety adverse events. The overall attack COVID-19 attack rate from 20/6/2020-27/9/2020 among registered nurses was 3.4% (102/2994): ICUs 2.2% (3/138), Geriatric wards 13.2% (26/197), and COVID-19 Wards 18.3% (32/175). The COVID-19 attack rate among medical staff was: all junior medical staff 2.1% (24/932), senior medical staff 0.7% (4/607), aged care/rehabilitation 6.7% (2/30), and ICU all medical staff 8.6% (3/35).

**Conclusions:** The isolation hood was strongly endorsed by staff and patients, and post-study became part of standard ICU therapy. ICU nurse COVID-19 infection rates were low. ICU HCWs feel safer when treating patients with COVID-19 using an isolation hood.

## Introduction

Since December 2019,^1^ the SARS-CoV-2 virus has led to the coronavirus disease (COVID-19) pandemic.^2^ The World Health Organization indicates that approximately 14% of people with COVID-19 require hospitalization (with O_2_ support), and 5% require intensive care unit (ICU) admission.^2^ Controversy surrounds the degree to which SARS-CoV-2 virus is spread via more localized droplets versus more distantly spread aerosols.^3, 4^ There is ongoing concern about SARS-CoV-2 infectious spread to healthcare workers (HCWs), and particularly from aerosol generating procedures (AGPs) such as intubation/extubation, nebuliser therapies, high flow O_2_, and non-invasive ventilation.^5, 6^

During the COVID-19 Pandemic the prevention of HCW infections has largely focussed on personal protective equipment (PPE).^7^ Controlling ventilation to avoid source spread from infected patients is arguably as important than appropriate PPE.^8^ Negative Pressure isolation Rooms (Class N= NPRs) can provide greater ventilation control than open-plan rooms but are a limited resource.^9^ Hospital NPRs provide high flows (12 air changes/hour) and negative pressure to prevent the spread of pathogens beyond the room’s confines,^9^; however infectious spread to personnel *within* the room remains problematic. Personal ventilation devices to protect HCWs and other patients from respiratory infections have been explored during prior and current infectious diseases outbreaks, such as SARS (2003),^1^ and COVID-19.^10, 11^

We hypothesised that a novel patient isolation hood had the potential to reduce HCW COVID-19 infections. To test this hypothesis we conducted a prospective cohort study to evaluate the safety and comfort of prototype personal ventilation hoods in a clinical setting. We also prospectively captured the number of HCW COVID-19 infections in the health service and the ICU.

## Methods

We undertook a prospective, interventional study of 20 patients whose management included the use of a personal isolation hood. This hospital ethics approved study for a Therapeutic Goods Administration (TGA) listed isolation hood device was conducted from 20/6/2020-21/7/2020 in the two general Intensive care units (ICUs) and Emergency Departments (EDs) at a metropolitan healthcare service in Melbourne. Feedback from participants and staff was obtained via a structured questionnaire. Two independent data safety monitors provided stewardship of trial conduct and adverse event reporting. Further details about the personal isolation *McMonty* Hood^12^ and the TGA adapted^13^ The hospital Ethics Committee Adverse Events Reporting form are contained in Supplementary Appendix 1. We prospectively monitored routine de-identified COVID-19 data of our institution’s HCWs. A confirmed COVID-19 infection was defined as a positive SARS-CoV-2 test (see Supplementary Appendix 1 for SARS-CoV-2 tests used).

Eligible participants for this trial were adult patients (≥18 years), being cared for in the ED or ICU, who were COVID-19 suspected/confirmed, or any respiratory infection warranting droplet or airborne precautions. Patients were excluded if: aged <18 years, pregnant, delirious, history of dementia, claustrophobia, and patients at risk of injuring themselves or others.

Participants or their medical treatment decision maker (MTDM) provided consent for use of the personal isolation hood (Supplementary Appendix 2). Healthcare staff who cared for patients using the isolation hood were invited to complete a single anonymous staff questionnaire (Supplementary Appendix 3); consent was implied if the questionnaire was attempted. Participants were invited to complete a feedback questionnaire and consent was implied if this was attempted (Supplementary Appendix 3).

Participants were free to open the hood cover or discontinue in the trial at any time. Staff could also cease use of the hood at any time. Use of the hood ceased when a patient had: (i) been declared negative for COVID-19 or another respiratory infectious disease, (ii) completed at least 7 days of treatment and deemed clinically appropriate by staff to cease use, (iii) been discharged from the ICU or hospital, or (iv) withdrew from the study.

### Survey Design and Data Analysis

The *staff* questionnaire comprised 18 closed and two open questions. Four of the closed questions were Likert scale questions (a 10-point Likert scale, numbered 1-10), and 14 Yes/No questions. The questions assessed the staff’s perception of the device’s ability to prevent cross-contamination, its safety (construction, mobility) and its practicality (patient access, communication). The *patient* questionnaire comprised seven closed questions (five Likert scale, and two Yes/No questions), and two open questions. The questions assessed the patient’s perception of the isolation hood’s, comfort, safety and ability to reduce infectious spread to healthcare workers. Free text areas were available for all questions to allow further patient/staff commentary.

We deemed that each questionnaire required a minimum of 50% of questions to be answered for data inclusion. Questions with a Yes/No answer were deemed 1/0 points for positive and negative responses respectively. Questions with a scaled 1-10 answer had a value of 0 (negative response) for a value of 1-5, or 1 (positive response) for a value of 6-10. Questionnaires were deemed either overall favourable (50% or greater positively answered) or unfavourable (less than 50% positively answered). A 75% or greater favourable response rate across all patient and staff questionnaires was deemed favourable isolation hood endorsement. Questions with more than one answer or no answer provided were excluded.

### Healthcare worker SARS-CoV-2 Infection Data

Treatment for COVID-19 hospital patients occurred in the ED, ICU, and designated COVID-19 Wards. We obtained prospective data of the observed rates of ICU health care worker (HCW) compared with other hospital HCW COVID-19 infections (all de-identified). We calculated the proportion of registered nurses and medical staff who developed COVID-19 both for ICU staff, and other hospital staff working elsewhere within the health service. We did not distinguish between SARS-CoV-2 infections at work/home/elsewhere; only COVID-19 positivity. We considered that staff could become symptomatic or test positive for COVID-19 up to 14 days post-study involvement. HCW Data are reported from the 20/6/2020 – 27/9/2020 (date of last COVID-19 diagnosis at our institution).

## Results

### Patient Characteristics

Twelve patients with confirmed, and eight patients with suspected COVID-19 were enrolled from 20/6/2020-21/7/2020 (Figure 1). Nineteen patients were treated in the ICU and one in ED (only).

**Figure 1:**
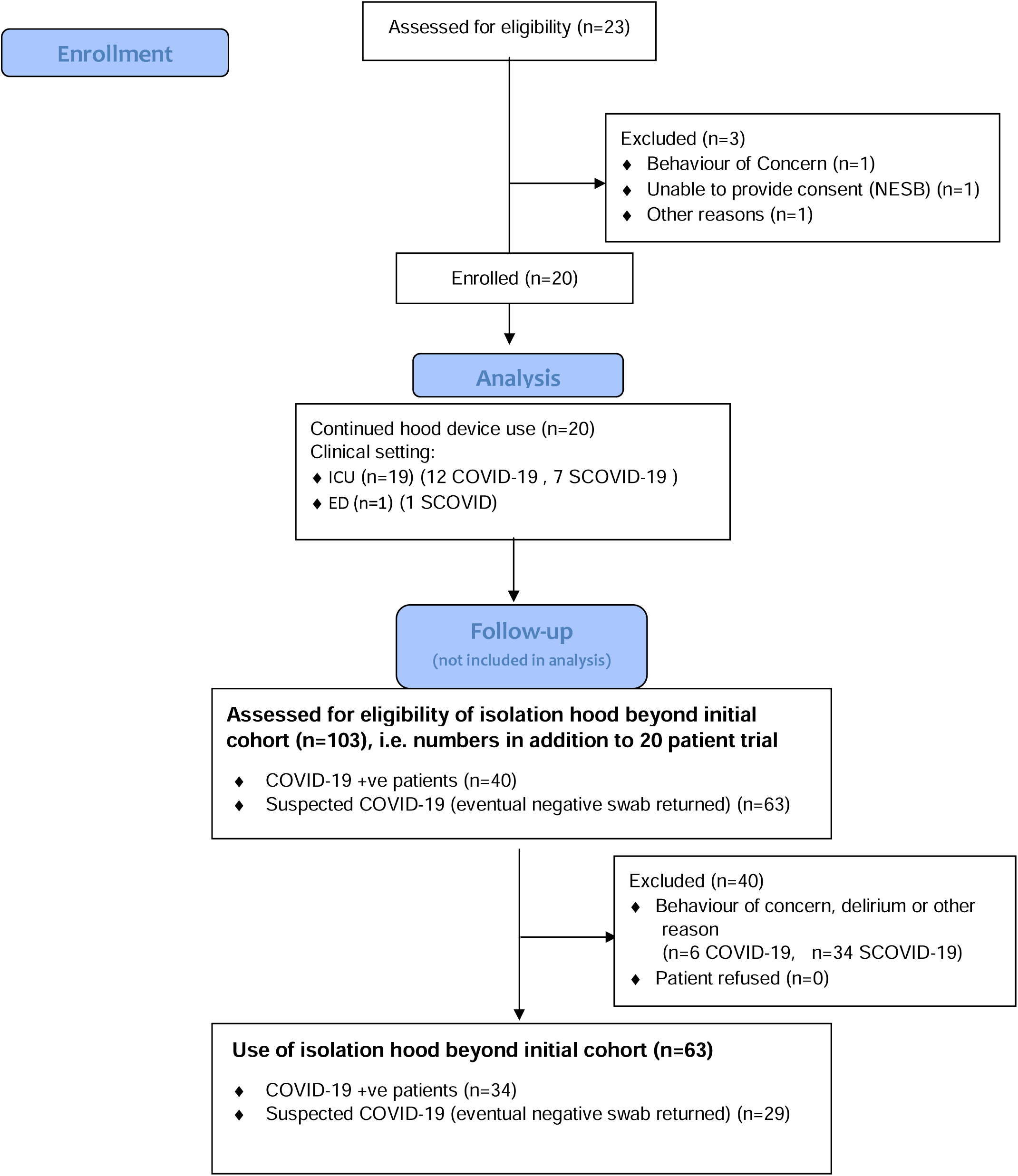
CONSORT Flow Diagram.

Eleven patients received invasive ventilation, five received non-invasive ventilation and two received nebuliser therapy (Table 1). The isolation hoods continued to be used beyond this study (i.e. after all questionnaires were completed on the 21/7/2020). After the 20-patient trial, 103 patients were screened for use of the isolation hood (40 excluded, all due to behaviour of concern, and delirium). A total of 34 COVID-19 patients and 29 suspected COVID-19 patients received the isolation hoods post-study, i.e. from the 22/7/2020 to 27/9/2020 (last date a HCW at our institution was diagnosed with COVID-19).

**Table 1:**
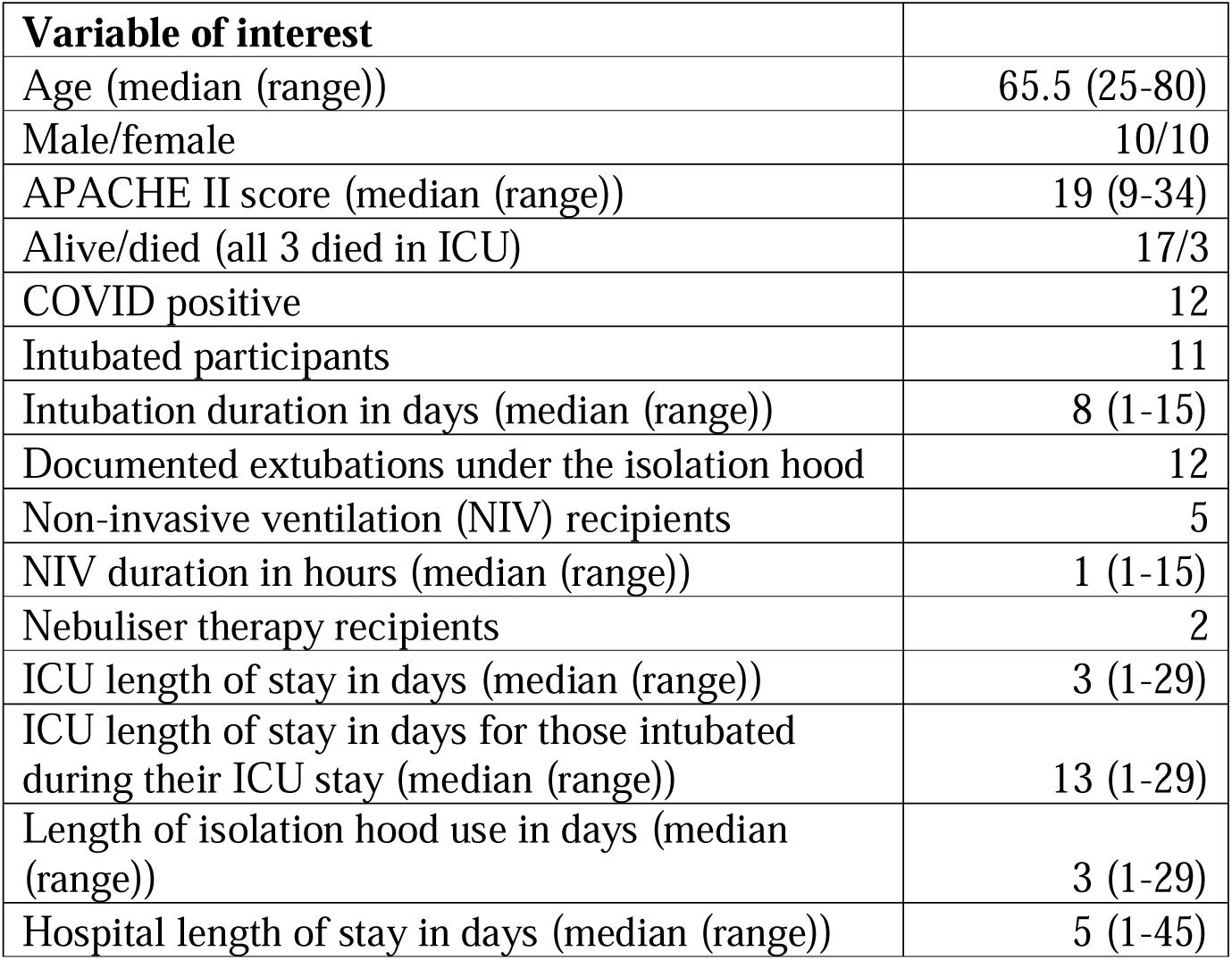
Patient Demographics and Therapies.

| **Variable of interest** |  |
| --- | --- |
| Age (median (range)) | 65.5 (25-80) |
| Male/female | 10/10 |
| APACHE II score (median (range)) | 19 (9-34) |
| Alive/died (all 3 died in ICU) | 17/3 |
| COVID positive | 12 |
| Intubated participants | 11 |
| Intubation duration in days (median (range)) | 8 (1-15) |
| Documented extubations under the isolation hood | 12 |
| Non-invasive ventilation (NIV) recipients | 5 |
| NIV duration in hours (median (range)) | 1 (1-15) |
| Nebuliser therapy recipients | 2 |
| ICU length of stay in days (median (range)) | 3 (1-29) |
| ICU length of stay in days for those intubated during their ICU stay (median (range)) | 13 (1-29) |
| Length of isolation hood use in days (median (range)) | 3 (1-29) |
| Hospital length of stay in days (median (range)) | 5 (1-45) |

### Staff Surveys

Of the 64 individual staff surveys, 60 (94%) were eligible (>=50% of questions answered), and all 60 surveys were overall favourable (>75% questions answered in favour of the isolation hood); see Table 2. The final two questions (infection control/hood cleaning) for the staff questionnaire remained unanswered as cleaning of the hood was undertaken by ICU research staff rather than ICU nurses for the study, and post-study by hospital cleaning staff. Staff were unanimous in: preferring to use the isolation hood (Q1), perceiving the hood as safe (Q2), understanding how the hood worked to reduce cross contamination (Q4), and found the hood to be in good working order (Q7). The isolation hood was perceived as robust and mobile by 53/57 (93%, Q.9), and reduced their chance of being infected with COVID-19 (Q10, 56/60= 93%), and that they felt comfortable administering AGPs to patients (Q15, 21/23 =91%). A relative majority (Q3, 41/60 =68%) of staff agreed the hood did not interfere with patient care. Staff also made free text comments which are further detailed in Supplementary Appendix 4.

**Table 2.** Staff Survey: Responses to Questions.

| Question | Theme | Number of favourable responses/ total responses (%) |
| --- | --- | --- |
| 1 | Prefer the hood compared to standard care | 56/56 (100%) |
| 2 | Safety of using device | 60/60 (100%) |
| 3 | Interference with patient care | 41/60 (68%) |
| 4 | Understanding of how the device protected healthcare workers | 60/60 (100%) |
| 7 | Physical condition of the device at the start of the shift | 58/58 (100%) |
| 8 | Ability to rapidly stow/remove the device in emergencies | 40/56 (71%) |
| 9 | Robustness and mobility of the device | 53/57 (93%) |
| 10 | Perceived reduced probability of contracting COVID-19 when using the device | 56/60 (93%) |
| 11 | Effectiveness of instructions printed on device | 48/54 (89%) |
| 12 | Location of patient-access points | 45/52 (87%) |
| 13 | Quality of device components | 52/59 (88%) |
| 14 | Accumulation of moisture/ exhalation/sputum | 56/58 (97%) |
| 15 | Staff comfort performing AGPs with the device | 21/23 (91%) |
| 16 | Ability of device to prevent cross-contamination | 57/58 (98%) |
| 17 | Isolation hood prevents communication | 15/22 (68%) |
| 18 | The patient appeared comfortable | 17/22 (77%) |

Table 3 shows the nurse-reported proportion of that nursing shift that the isolation hood was used in the ‘hood down, fan on’ configuration (versus hood up, fan off *or* on). The reported proportions are exclusive of the time the hood was opened for necessary clinical reasons, e.g. for patient turning. 84 logs were recorded; 51/84 (61%) of staff reported used the hood down/fan on for more than 75% of the shift. Common reported reasons to not use the hood in this configuration were due to patient concerns and requests for a break from the isolation hood.

**Table 3.** Staff reported use of isolation hood in the ‘hood down, fan on’ configuration across a nursing shift.

| Proportion of shift when hood was down, fan on | Number of responses (%) |
| --- | --- |
| 0-24% of shift | 12 (14%) |
| 25-49% of shift | 6 (7%) |
| 50-74% of shift | 15 (18%) |
| 75-100% of shift | 51 (61%) |

### Patient Responses

Eight participants (8/20, 40%) completed the questionnaire. Three patients died during their ICU admission, and nine patients did not complete the questionnaire. Seven of eight patient surveys were favourable (>75% questions answered in favour of the isolation hood). All eight patients who completed the patient questionnaire indicated that they thought that the isolation hood: helped prevent COVID-19 cross-infection, was safe to use, and comfortable. Most (7/8=88%) patients thought that the hood was easy to open by them, and that the temperature and humidity was comfortable (6/8=75%). Patients agreed less strongly that the hood provided temperature and humidity comfort (5/8= 62.5%), and that they could communicate adequately whilst inside the hood’s canopy (5/8= 62.5%). Patients made free text comments about three questions which are given in Supplementary Appendix 4.

### Adverse events

No patient related safety adverse events were reported. All adverse events were technical concerns related to the isolation hood’s design or operation. The data safety monitors received two near incidents and nine non-incidents. Additional details of the non–incidents and rectification required are presented in Supplementary Appendix 4. The final non-incident involved an audible alarm that confirmed the fan was on (an additional feature to remind staff the fan should be on while the hood was down). The alarm remained on continuously, necessitating its removal. New prototypes were fitted with a light at front of the hood to indicate that fan was on.

### COVID-19 HCW Infections

All HCW COVID-19 infection data are from 20/6/2020 until 27/9/2020. The overall attack COVID-19 attack (infection) rate among registered nurses (RNs) was 3.4% (102/2994): ICUs 2.2% (3/138), Emergency Departments 3.2% (11/366), Surgical wards 1.2% (3/252), Geriatric Wards 13.2% (26/197), and COVID-19 Wards 18.3% (32/175). The COVID-19 attack rate among medical staff was: all junior medical staff 2.1% (24/932), senior medical staff 0.7% (4/607), anaesthetists 1.9% (2/104), aged care/rehabilitation 6.7% (2/30), and ICU all medical staff 8.6% (3/35).

## Discussion

We report the first clinical evaluation of a novel patient isolation hood used during the 2020 COVID-19 pandemic. We found extremely high levels of patient and staff satisfaction: post-study the isolation hoods became part of standard ICU therapy. No device related patient adverse events were reported. Several technical adverse events were reported, which will inform future device design and development. No single question was answered negatively about the isolation hood. Improved safety (from COVID-19 cross-infection) was the most common and pronounced reason why staff liked the isolation hood. Patients thought that the isolation hood helped prevent COVID-19 cross-infection, was safe and comfortable. Attack rates of ICU nurse COVID-19 infections were lower than other nursing staff groups.

Aerosol generating procedures such as non-invasive ventilation, endotracheal extubation, and nebuliser therapy were able to be delivered in our open planned ICUs in the presence of the isolation hoods without recourse to transports to/from a negative pressure room. It is unclear, however, whether the use of isolation hoods affected clinical outcomes. Prior studies regarding the use of isolation hoods during pandemics, including COVID-19.^10-12,14^ exist. Adir et al recently reported the positive responses of a survey that included nine staff members. To our knowledge, this is the only study that reports staff’s and patients’ views about the use of an isolation hood during COVID-19.^11^

No major adverse events occurred during the study, and as we proceeded new isolation hood prototypes were modified to have a lower centre of gravity, a fan-on light, and more robust plastic canopies. The least-strongly supported theme for the isolation hood of the staff survey related to communication and interference with patient care. Negative patient comments about the hood mainly related to fan noise. This feedback will inform future design.

Of the total number of coronavirus (COVID-19) cases in healthcare workers in Victoria, Australia (3,574 as of November, 20^th^ 2020), 73% of cases were acquired in a healthcare setting.^15^ Observational data of HCW infections in Victoria indicate that COVID-19 infections in ICU staff are less common than among HCWs on COVID wards, hospital aged care wards and aged care workers in residential aged care facilities (Marion Kainer, personal communications). This study was conducted in open planned ICUs with only one single negatively pressured room used solely for intubations/bronchoscopies/tracheostomies.

The ICU nurse COVID-19 attack rates were the second lowest of any hospital nursing group. Potential factors other than the use of a patient isolation hood that could influence this observation include: patients with COVID 19 admitted to ICU are typically a week from symptom onset and potentially less infectious; improved ICU nurse to patient ratios; the use of invasive mechanical ventilation with exhaled gas HEPA filtration; ICU nurse PPE training, and different room ventilation. In Victoria, Australia, however, hospital building guidelines state that air exchange rates must be six per hour for ICUs *and* for all medical/surgical ward areas.^9^ If these guidelines are implemented, the rate of air exchange is not a reason for different observed COVID-19 HCW infection rates. The ICU medical staff COVID 19 attack rate was numerically higher than other medical staff groups. ICU medical staff routinely performed endotracheal intubation (where the isolation hood was unable to be used).

Our study has limitations. By its nature it was unblinded, and the single health centre sample size was small. We did not distinguish between nurse/doctor/other questionnaire responses, or conduct detailed investigation of infection control procedures. Further bias may have arisen due to ICU staff and ICU researcher familiarity. We did not measure viral loads within/exterior to the isolation hoods. We had no ability to adjust for potential confounders. While the observed attack rate for ICU registered nurses was low it remains uncertain if the isolation hood reduces HCW COVID-19 infections: no causal inference may be drawn.

This study complements our pre-clinical assessment of the isolation hood’s efficacy of limiting aerosol spread.^12^ The results of this study support the conduct of translational research and implementation studies of the isolation hood in other hospital areas and other jurisdictions. This study provides evidence of the safety and comfort of an isolation hood as part of routine treatment of patients with COVID-19. There was a high rate of acceptance by patients and staff. It is apparent that HCWs feel safer when treating patients using a personal ventilation hood. It is plausible the isolation hood reduces COVID-19 HCW infections. Additional studies to define the role of this device are indicated.

## Data Availability

Data shall be made available by the authors where appropriate data requests are made.

## Acknowledgements

We thank the Western Health (WH) ICU research team: Rebecca McEldrew, Miriam Towns, and Rebecca Morgan for data collection, WH ICU and ED staff. We also thank the assistance of WH Research Manager Bill Karanatsios for TGA submission and notification etc. We thank the WH Infection Prevention team led by Richard Bartolo for input to the care and cleaning of the isolation hoods, patient consumer representatives John Ward and Grant Carroll for input on questionnaires. Assistance from the University of Melbourne Department of Mechanical Engineering (Kevin Kevin, Max Rounds, Geoff Duke) in the isolation hood prototype developments and improvements was integral. We also thank the University of Melbourne and The Western Health Foundation for funding to construct the first 17 isolation hoods as used in this clinical trial.

## Notes

### Competing Interest Statement

Conflicts Of Interest: A patent has been filed for the personal ventilation hood by the University of Melbourne/Western Health. The lead authors (Forbes McGain, Jason Monty) were the leads in this patent application. There are no other conflicts of interest.
All other authors have no conflicts of interest.

### Clinical Trial

Australian and New Zealand Clinical Trials Registry Number:ACTRN12620000500954p
World Health Organization Universal Trial Number: U1111-1250-6657

### Funding Statement

Funding: funding was obtained for this study from the Western Health Foundation
and the University of Melbourne to pay for the construction of the first 17 isolation
hoods.

### Author Declarations

HREC Reference Number: HREC/63694/MH-2020 Melbourne Health Site Reference Number: 2020.129 Project Title: A personalised ventilation hood for Covid-19: A phase 1 study of a new device.

