## Supplemental Appendices 1-4 for "A Prospective Clinical Evaluation of a Patient Isolation Hood During the COVID-19 Pandemic"

### Supplementary Appendices (1-4).

### Supplementary Appendix 1: Further Details of Methods and Adverse Events /Technical Complaints Reporting

**SARS-CoV-2 Testing.**

Four different tests were used during the study period to diagnose SARS-CoV-2:

1. Cobas SARS-COV-2, Roche Diagnostics, Mannheim, Germany,

2. Allplex SARS-CoV-2 Assay, Seegene Inc., Seoul, S. Korea

3. Wuhan CoV E –gene, Tib-Molbiol, Berlin, Germany

4. BGI (Beijing Genomics Institute) SARS-CoV-2BGI PathoGenesis Pharmaceutical Technology Co., Beijing, China.

**The Isolation Hood.**

The *McMonty* isolation hood^1^ comprises a mobile, transparent pram canopy enclosure, a fan, and an H13 high efficiency particulate air (HEPA) filter. The fan draws air from inside the hood, through piping, to the filter. The entire unit is reusable; i.e. cleaned following each patient use. Droplet spread is contained by the canopy, whilst aerosol spread is contained by the enclosure’s negative pressure induced by the fan (100 air changes/hour) drawing air away from the healthcare worker. Importantly, the isolation hood is an *open* system, allowing airflow from around the patient towards the fan/filter at the canopy’s rear.

**Safety Data and Adverse Events Reporting**

External, independent safety monitors (one clinician, one engineer) formed the Data and Safety Monitoring Board (DSMB). We reported all incidents/near incidents/non-incidents to the DSMB as they arose. A planned interim safety report was conducted after 5 participants had been recruited: a 75% favourable response rate was prospectively defined as being necessary for study continuation.

**Adverse events definitions:** there were three classifications of incidents. **Incident:** technical complaint directly/indirectly led to the death or serious deterioration of health of the patient subject or others. **Near Incident:** technical complaint could potentially lead to the death or serious deterioration of health (=serious injury) of the subject or others. **Non Incident**: technical complaint did not and could not lead to the death or serious deterioration of health (=serious injury) of the subject or others.

**Technical Complaints and Adverse Events Form**

**(Names have been removed for manuscript de-identification purposes)**

**Section 1: To be completed by the initiator of the complaint**

| To: | Medical safety monitors – | **From:** | | **Name of staff making the complaint:**  E-mail: |
| --- | --- | --- | --- | --- |
| Patient Study No: | |  | | |
| Study Manager: | | | | |
| This technical complaint is related to the (check all that apply) | | | ☐ medical device  ☐ other ________________________________ | |
| This technical complaint is associated with a/an | | |  | |
| Incident: This technical complaint directly/indirectly led to the death or serious deterioration of health (=serious injury) of the patient subject or others, e.g. staff.  Near Incident: This technical complaint could potentially lead to the death or serious deterioration of health (=serious injury) of the subject or others, e.g. staff.  Non Incident: This technical complaint did not and could not lead to the death or serious deterioration of health (=serious injury) of the subject or others, e.g. staff. | | | ☐ Incident  ☐ Near Incident  ☐ Non Incident | |
| **Description of Complaint** (specify exact information about the study medical device, and who discovered or reported the problem):  ***As examples: was there a malfunction of the device?, Did the patient become distressed by the McMonty hood?, Did people forget to turn the fan on when the hood was down?, Was it difficult to move the McMonty?, etc.).*** | | | | |
| Additional information attached: Yes ☐ No ☐  If yes, please specify : | | | | |
| Date:__________ Name: ________________________ Signature:______________________ | | | | |

**Section 2: To be completed by an Isolation Hood Trial Investigator**

| Date: | Form No: | |
| --- | --- | --- |
| Recipient of Complaint /Name/Signature : | | |
| Initial classification:  *Adverse device effect (ADE).* Adverse event related to the use of the investigational product.  *Adverse event (AE).* Any untoward medical occurrence, unintended disease or injury, or untoward clinical signs in subjects, users or other persons, whether or not related to the investigational medical device.  *Serious adverse effect (SAE)*. Any adverse effect that results in any of the following outcomes: death, a life-threatening adverse effect, inpatient hospitalization or prolongation of existing hospitalization, a persistent or injury or permanent impairment to a body structure or a body function.    *Unanticipated serious adverse device effect (SADE).* Serious adverse device effect which by its nature, incidence, severity or outcome has not been identified in the current version of the risk analysis report. | Adverse Device Effect  Adverse Event:  Serious Adverse Event:  Unanticipated Serious Adverse Device Effect: | ☐  ☐  ☐  ☐ |
| Date:__________ Name: ________________________ Signature:______________________ | | |

**Section 3:	To be completed by Medical Safety Monitor: Final evaluation**

| To: | Name:  Address: | **From:** |
| --- | --- | --- |
| Complaint System No.: | | Final classification:  Category: |
| Final assessment: ☐ Confirmed quality defect  ☐ unconfirmed quality defect  ☐ unconfirmed quality defect but plausible | | |
| Response to Complaint: | | |
| Additional information attached: Yes ☐ No ☐  If yes, please specify : | | |
| Date:__________ Name: ________________________ Signature:______________________ | | |

1 McGain F, Humphries RS, Lee JH, et al. Aerosol generation related to respiratory interventions and the effectiveness of a personal ventilation hood. *Critical Care and Resuscitation* 2020; **22**: 212-20.

**Supplementary Appendix 2: Staff Questionnaire for the McMonty Patient Isolation Hood**

Question 1. **For future patient care, would you prefer to use the McMonty hood for patients with suspected or confirmed COVID-19, instead of standard care/no hood?**
Question 2. **Did you feel that the McMonty hood was safe to use?**
Question 3. **Did the McMonty hood interfere with patient care?**
Question 4. **Did you understand how the McMonty hood worked to attempt to reduce COVID-19 cross contamination by droplet spread and aerosol generation?**
Question 5. **What features did you like about the McMonty hood? (Utility, safety, mobility, size, shape, etc.)**
Question 6. **What features did you dislike about the McMonty hood? (Utility, safety, mobility, size, shape, etc.)**

Question 7. **Please check the physical state of the plastic hood for rips/tears prior to use/reuse. Please indicate if the state of the plastic hood or the frame, wheels or fan were in good working order.**
Question 8. **Was the McMonty hood readily mobile on its wheels? Did you feel that you could rapidly move the hood out of the way in case of rapid patient deterioration (for example, preparation for intubation)?**
Question 9. **Was the actual plastic hood with the underlying ribs/scaffolding robust and mobile enough to your suiting?**
Question 10. **After using the McMonty hood did you feel less likely to contract COVID-19 from the patient you were caring for?**
Question 11. **Did the “HOOD DOWN = FAN ON” sign on the front of the plastic hood remind you to turn the fan ON when the hood was DOWN?**
Question 12. **Was the position of the metal side arms or location of the side flaps in the plastic hood suitable for routine patient care?**
Question 13. **Did the fan or any other part of the McMonty hood fail/not live up to expectations at any stage?**
Question 14. **Did you have to clean the inside of the hood with alcohol based disinfectant or detergent due to excessive moisture/exhalations/sputum etc. at any stage?**

**If yes, how often did you clean the interior of the hood per day?**

Question 15. **Were any aerosol generating procedures (nebulisation, extubation, high flow nasal O_2_ or BiPAP) administered to your suspected or confirmed COVID-19 patient at any stage with the McMonty hood on?**

**If yes, please write which therapies?**

**If yes, did you feel comfortable using the McMonty hood for these aerosol generating procedures?**

Question 16. **Overall, how would you rate the McMonty hood as a device to prevent cross contamination of staff and other patients with COVID-19?**
Question 17. **For the awake patient did the hood prevent communication?**
Question 18. **Did the awake patient indicate either verbally or non-verbally that they felt uncomfortable within the hood?**
Question 19. **During the dismantling of the hood, did you feel this process was achieved safely, in a way that would minimise potential contamination?**
Question 20. **Did you send the plastic hood off for cleaning?**

**If yes, was the packaging of the hood into the ‘duffel bag’ simple and straightforward?**

**Supplementary Appendix 2: Patient Questionnaire for the McMonty Isolation Hood Study**

**Question 1.**	**Did you feel that the McMonty hood helped prevent any infectious spread that you may have to other people?**

**Question 2.** **On a scale of 1-10 did you have any concerns about the physical structure of the McMonty hood and your physical safety within it?**

**Question 3.** **On a scale of 1-10, how comfortable did you feel by the enclosed space?**

**Question 4.** **Did you feel the temperature and humidity inside the hood was comfortable?**

**Question 5.** **Did you find the hood fan too noisy?**

**Question 6.** **Did the hood reduce your ability to communicate with staff to the point where you could not be understood?**

**If yes, how was your communication affected?**

**Question 7.** **If you wanted to open the hood could you do so easily?**

**Question 8. What were the positive features (if any) you liked about the McMonty hood?**

**Question 9. What were the negative features (if any) you disliked about the McMonty hood?**

### Supplementary Appendix 4: Results for the Questionnaire Free Text Comments, and Adverse Events

### Free Text Comments by Staff

Eleven staff wrote that they would rather have the isolation hood present than not when caring for patients with COVID-19 (Q.1), with no negative comments. All eleven free text answers to Q.1 indicated that the hood made staff feel safe, and provided an additional layer of protection beyond PPE, particularly for non-invasive ventilation and nebulisation. Two staff indicated that it made them “feel less anxious about working on the front line.”

Most staff noted that it was: easy to lift the plastic canopy out of the way, mobile, and in an actual experienced emergency such mobility allowed for prompt removal. Two staff thought that the isolation hood was difficult for short statured HCWs, two staff noted that one had to be more mindful of the endotracheal tube position, and a further two found patient oral care more difficult. Q.5. asked about what features were liked about the isolation hood with 27 responses of which 10 found it easy to use, eight liked its safety aspect, and seven its transparency.

Nine staff made comments about what they disliked about the isolation hood: four noted the device’s bulkiness, wheel brakes difficult (two staff), plastic canopy slits for access could be positioned differently (one staff), position the hood in a half-way position for ease-of-access (one), and height too challenging (one). Staff felt comfortable undertaking AGPs when the hood was in use [21/23 (91%)]. A minority of staff answered questions 17 (Isolation hood prevents communication) and 18 (the patient appeared comfortable), although the majority wrote favourably. Because the ICU researchers undertook much of the cleaning during the study (followed by cleaning staff thereafter), questions 19 and 20 about cleaning remained unanswered.

### Free Text Comments by Patients

Positive comments about the hood were: “I liked how I can still see activity happening around me”, “Normally I have to go in an isolation room when I come to hospital and it’s very lonely. You can see nobody for hours.”, “This hood lets me still be seen by staff and I don’t feel forgotten about.” and, “I felt that it helped to stop the spread of infectious diseases.

Negative comments about the hood were: noise: “Just have to get staff to speak up, I just spoke up a little more, and it was ok”, and “I had to yell and try to open the hood, which I couldn’t do easily”, lighting: “The overhead lights bounced off the plastic a little bit, depending on the angle of the plastic, but not troublesome”, and temperature: “I felt a bit trapped and too hot.”

### Details of Adverse Events /Technical Complaints Reporting

There were two near-incidents: these both involved the hood tipping forward, (once with the hood’s brakes disengaged). Later isolation hood prototypes had extensions to the hood’s base in order to prevent forward tilting.

There were nine non-incidents: four tears of the plastic canopy, three loose screws, a single episode where the fan housing loosened, and a fan alarm malfunction. The final non-incident involved an audible alarm that confirmed the fan was on (an additional feature to remind staff the fan should be on while the hood was down). The alarm remained on continuously, necessitating its removal. New prototypes were fitted with a light at front of the hood to indicate that fan was on.
